# Organization of Primary Health Care in pandemics: a rapid systematic review of the literature in times of COVID-19

**DOI:** 10.1101/2020.07.05.20146811

**Authors:** Thiago Dias Sarti, Welington Serra Lazarini, Leonardo Ferreira Fontenelle, Ana Paula Santana Coelho Almeida

## Abstract

The world is experiencing one of the greatest public health emergencies in history with the global spread of COVID-19. Health systems, including Primary Health Care (PHC) services, are pillars of pandemic coping strategies, and there are important gaps in the literature on the best ways to organize PHC in health crisis scenarios such as the one currently experienced. Given the urgency of responses, we performed a rapid systematic literature review on MEDLINE (via PubMed), EMBASE and LILACS (via VHL), in order to analyze empirical studies on the effectiveness of PHC organization strategies in the context of epidemics to improve access and reduce morbidity and mortality. We selected seven articles, which studied the responses to different epidemics in different parts of the world. In terms of access, the studies suggest positive results with the adoption of adjustments of work processes of the teams and the structure of the services, combined with diversification of actions (including call center), adequate provision of inputs and personal protective equipment, adequate action plans and communication strategies, and effective integration with public health services and other levels of care. No study analyzed population morbidity and mortality. The included studies suggest also that community-oriented PHC is more effective in crisis scenarios, indicating the necessity of strengthening of the Family Health Strategy in the Brazilian context.

## Introduction

Major epidemics have marked history, causing significant social, cultural, political, economic, and health impacts ^1^. The past few decades have seen the emergence of new pathological agents that have spread fear in most parts of the world. These are the cases of SARS (Severe Acute Respiratory Syndrome), beginning in 2002 in China, which affected approximately 8 thousand people in less than 1 year, and MERS (Middle East Respiratory Syndrome), which reached less than a thousand people in the two years after its emergence in Saudi Arabia, in 2012 ^2^. These figures illustrate the unprecedented dimension that a new pandemic (also caused by a coronavirus, SARS-CoV-2), gained in approximately 5 months of global dissemination, from its epicenter in Hubei province, China, reaching approximately 3.5 million infected and 250,000 deaths at the beginning of May 2020 ^3^.

The magnitude of COVID-19 makes it one of the greatest humanitarian crises of the last few centuries and a public health emergency, understood here as an extraordinary occurrence, which puts other States at risk and requires coordinated and robust international strategies for disease prevention and control ^4,5^. The enormous speed of contagion and spread between countries and regions, combined with the severity of the disease, pressures for decisions that affect the lives of the entire population to be taken quickly and with high degrees of uncertainty as to their effectiveness ^6^. The sciences are used to point out the best strategies for social distancing and individual protection, developing tests to identify cases, assessing the effects of antiviral drugs and potential vaccines, aligning clinical protocols, and organizing health systems. But in each aspect of these, the novelty of the pandemic and the current political configurations challenge the current methodologies of production, dissemination of knowledge, and decision making at different levels of social organization.

The organization of health systems and services in the face of a pandemic is a pillar of any emergency plan, whether local, national, or international. In general, systems that have robust Primary Health Care (PHC) are more effective, efficient, safe, equitable and sustainable, since this level of care is considered central in all types of response to acute or chronic health problems ^7,8^. In this sense, it is possible to state that PHC should be considered one of the main strategies for coping with a public health emergency, including COVID-19. Ease of access to a population already known by health teams trained to provide comprehensive care in a socially oriented manner and in coordination with the other levels of care make PHC an essential tool in both the short and the medium and long term coping with the various waves of a public health emergency ^9,10^.

However, there is little scientific evidence on the most effective strategies for organizing PHC services; most available evidence refers to the hospital environment ^11^. The demand for timely responses to an epidemic leads us to think pragmatically about the format of local healthcare flows to serve an increasing number of sick people. However, incorporating knowledge produced in the context of an epidemic can contribute to a more rational decision making, resulting in an adequate use of supplies and equipment (which are difficult to obtain in times of high demand at sustainable costs) and less population morbidity and mortality.

Therefore, this study aimed to carry out a rapid systematic review of the literature that analyzes the effectiveness of PHC organization strategies in the context of epidemics.

## Methods

In this rapid systematic review ^12^, we included original observational or experimental quantitative as well as qualitative empirical articles that have been peer-reviewed and published in indexed scientific journals, with no date limit, in English, Portuguese or Spanish. Studies should focus on analyzing the organization and structuring of PHC services to face a critical public health situation, characterized as an epidemic or pandemic. The articles also needed to define as an outcome the facilitation of the population’s access to health care or population morbidity and mortality. We excluded studies that did not address PHC, did not bring original empirical data, that did not define a desired outcome of the intervention, or had not been carried out in the context of an acute event with a significant impact on public health. Opinion articles, comments, essays, letters, editorials, experience reports, and reviews were also excluded.

Systematic searches for articles were made on April 1, 2020 by one of the authors (APSC) on MEDLINE (via PubMed), EMBASE, and LILACS (via VHL). No procedures were established for searching the gray literature. The search strategy used was (“primary health care” OR “primary care” OR “community health service”) AND (“Patient Care Management” OR “health care management” OR “Organizational Innovation” OR “Personnel Management” OR “health care personnel management” OR “Professional Practice” OR “Program Development” OR “Quality of Health Care” OR “Total Quality Management”) AND (“Disease Outbreaks” OR epidemic OR “Disaster Medicine”). This search strategy was built from MeSH (Medical Subject Headings) and Emtree (Embase subject headings) descriptors, adapting the searches to the MEDLINE and EMBASE databases, respectively; the MEDLINE search strategy was reused in LILACS.

The search results were condensed and transferred to EndNote X7®. Duplicate articles were excluded. Two researchers (TDS and APSC) independently screened titles, abstracts, and full articles for analysis of the work’s eligibility. Disagreement was resolved by consensus and, if necessary, by a third researcher (LFF).

Data extraction was performed by two authors (TDS and WSL) with an instrument designed for the purposes of this research. The articles were characterized based on the following information: authors, year of publication, title, place and year of realization, and the journal publishing the study; objective, methodological design, study scenario, sampling, and data analysis techniques; type of acute event faced (eg H1N1, SARS-CoV, MERS-CoV); outcome or improvement aspect of the analyzed service; main results of the study.

The objective and exploratory character of this review and the methodological diversity of the studies led us to analyze the results in a narrative manner, identifying aspects of the organization and structuring of PHC services that result in handling the crisis situation, considering that this review was planned in order to serve as a basis for reflections and decision-making by managers and health professionals in their workplaces. In this sense, we chose not to carry out an in-depth analysis of the quality and potential of study biases individually. The potential impact of articles on the reality of health systems and global limitations of the scope and methodology of the studies were analyzed.

The research was reported following the PRISMA (Preferred Reporting Items for Systematic Reviews and Meta-Analyzes) recommendations ^13^, adapted as needed to the rapid systematic review format. The research was also registered in the PROSPERO (the international prospective register of systematic reviews) database with the code CRD42020178310, after ensuring there were no ongoing reviews with the same scope.

## Results

The search strategy resulted in 2473 articles, highlighting the absence of research identified in LILACS, which may denote problems of indexing Latin American journals or some invisibility of the topic in this region. The screening of titles, abstracts, and full texts resulted in the selection of seven articles ^14–20^ for the narrative synthesis presented here (Figure 1).

**Figure 1.**
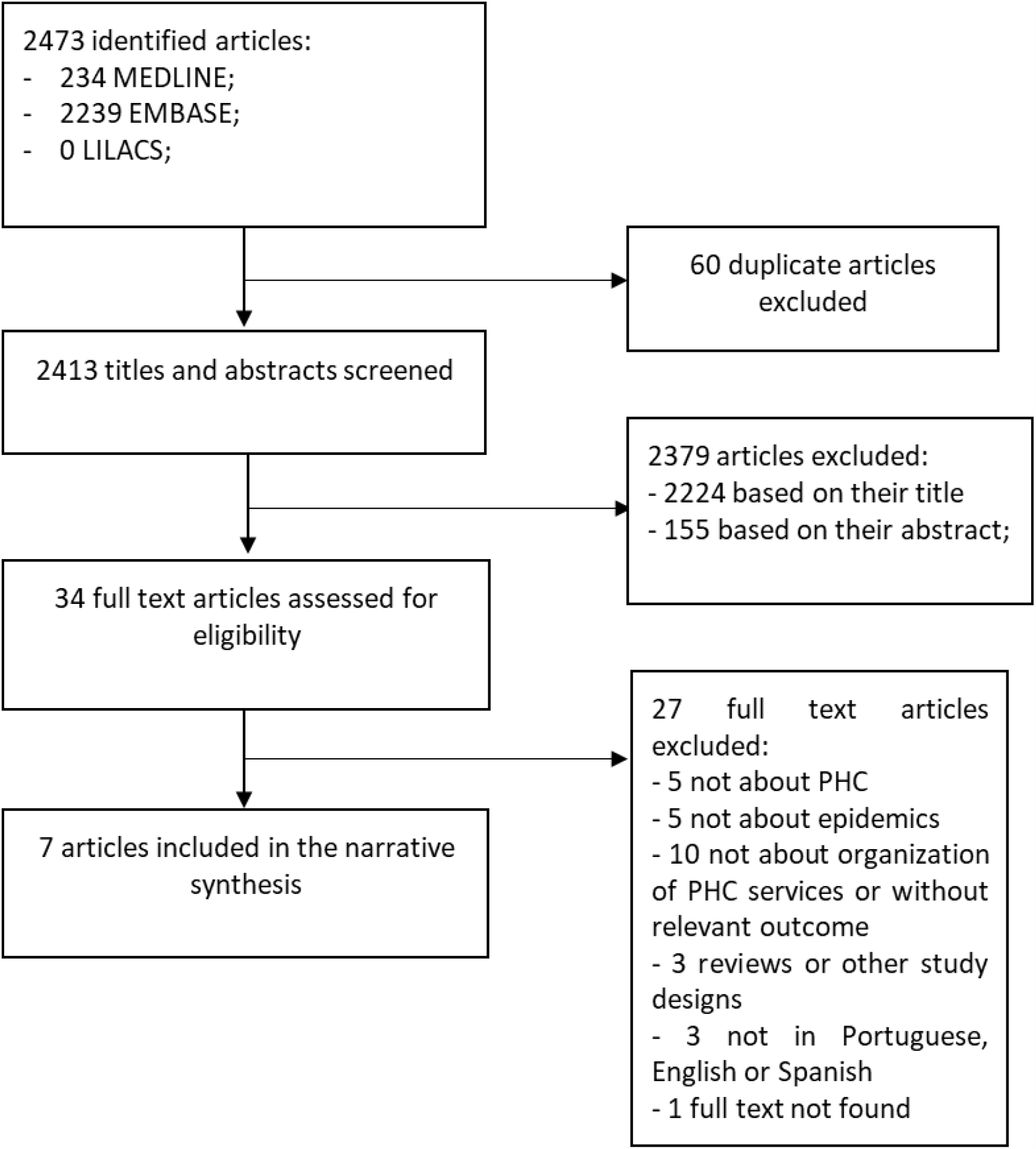
Flowchart of study selection.

Throughout the screening of the articles, we identified many studies that addressed chronic conditions of illness, problematic use of opioids, infectious diseases, mass vaccination campaigns, description of epidemiological surveillance systems, and studies focused on the hospital environment. It is interesting to observe how part of the academic literature links the problem of illness due to chronic conditions to the concepts of epidemic and even pandemic, denoting the significant impact that these diseases present in the world today, especially in more developed countries, and potentially a publication bias. In addition, there were numerous experience reports, opinion texts, theoretical models and protocols for action, government positions, and descriptions of the role of different professional categories and managers in a situation of public health crisis secondary to a major epidemic.

All seven articles were based on qualitative research methods, with two ^17,20^ triangulating such methods with administrative data or a survey with probabilistic sampling (Table 1). They aimed to analyze the effects of different epidemics on the population, health professionals and the organization of health services, and sought to understand the perspectives of different actors directly or indirectly involved with the health system, varying from managers and stakeholders linked to corporate organizations and non-governmental organizations to health professionals and community members.

**Table 1.** Characteristics of the studies.

| **Author,**<br>**year** | **Epidemic** | **Local** | **Undertake**<br>**n in year** | **Design** | **Subjects** |
| --- | --- | --- | --- | --- | --- |
| Bergeron et al., 2006<sup>14</sup> | SARS | Canada | 2004* | Postal survey with probabilistic sample including to open questions; thematic analysis. | Community nurses |
| Bocquet et al., 2010<sup>15</sup> | H1N1 | Australia | 2009 | Purposive sampling in health centers with high volumes of influenza-like illness presentations; semi-structured interviews, thematic analysis. | Family physicians |
| Masotti et al., 2013<sup>16</sup> | H1N1 | Canada | 2009* | Semi-structured interview with key informants, with comparative case study. Nominal group technique via Internet. | Medical officers of health, family physicians |
| Miller et al., 2018<sup>17</sup> | Ebola | Guinea, Liberia e Sierra Leone | 2016 | In-depth interviews and focal groups; thematic analysis followed by triangulation with administrative data. | Stakeholders from local authorities, external multilateral agencies, professional organizations leaders, third sector, community health workers, community leaders, healthcare users |
| Phillips et al., 2007<sup>18</sup> | SARS | Australia | 2006 | Individual and group interviews and workshops; simulation with PHC workers of three models of healthcare delivery | Healthcare managers, nurses and family physicians, leaders of professional organizations and third sector stakeholders |
| Sarikaya, Erbaydar, 2007<sup>19</sup> | H5N1 | Turkey | 2006 | Semi-structured interviews with key informant; thematic analysis. | Healthcare managers, family physicians, nurses and midwives |
| Siekman et al., 2017<sup>20</sup> | Ebola | Liberia | 2015 | Focal groups with purposive sampling (gender, diversity of experience). Survey with probabilistic sampling. Interviews with key informants. Content analysis and triangulation with administrative data. | Community health workers |

\* Probable year, since authors didn't explicit it. PHC, primary health care. SARS, severe acute respiratory syndrome.

Based on these characteristics of the seven selected studies, which are quite heterogeneous with each other, we proceed to a narrative synthesis of the main findings that can bring relevant contributions to the structuring of responses from health systems worldwide (Table 2). We chose to group these results in some thematic groups, which we present below.

**Table 2.** Main findings of the included studies.

| **Article** | **Setting** | **Findings** |
| --- | --- | --- |
| Bergeron et al., 2006<sup>14</sup>. (SARS). | • Diverse healthcare services in Ontario, including public health, home care, Community Care Access Centres and clinics, among others.<br>• Canada was the country outside Asia most severely affected by SARS, and healthcare workers constituted two thirds of the probable cases. | • Pillars of effective confrontation of the epidemic: leadership; communication; resource allocation; recognition and support for the professional; participation of professionals in organizational decision-making; Health education.<br>• Effective communication between managers and healthcare workers and between them and users and the community must be a priority, with priority actions clearly defined.<br>• Organizational support is essential, meaning that managers are concerned with the safety of healthcare workers, respond to their concerns, and resolve conflicts.<br>• Guidelines and protocols for emergencies are fundamental.<br>• Main challenges: work overload; emotional impact (fear, stress, isolation, and frustration); lack of personal protective equipment. |
| Bocquet et al., 2010<sup>15</sup>. (H1N1). | • Physician-centered solo, group, and corporate PHC practices.<br>• Many clinics with a high proportion of users per doctor, offering access to vulnerable populations.<br>• Funding of clinics: public (bulk billing), private (fee-for-service), or mixed.<br>• Public health authorities support PHC, including the provision of PPE and standardization of clinical guidelines. | • The most effective clinics had stocks of PPE, strict professional hygiene and protection protocols, rapid action plans for epidemics, communication strategies, and underwent training in a timely manner.<br>• The structure of the clinic was decisive for the expansion of care for symptomatic patients and maintenance of care for non-affected users: alternative entrances to the service that could be destined for the affected, extra rooms designated for the exclusive care of the affected, and large waiting rooms.<br>• Some clinics have designated professionals (nurse or doctor) for the exclusive attention of symptomatic users, including administrative and epidemiological surveillance activities, but it was not possible to identify the results of this initiative.<br>• Communication between health authorities and clinics must be synchronized with the concrete demands of professionals, oriented towards clinical practice, specific to PHC, and easily accessible and understood.<br>• Specific clinics for the care of symptomatic patients were created near the hospital structure. They were considered important in the rear of both the PHC and the hospital, mainly during non-business hours.<br>• Smaller PHC clinics with problems with PPE stock or removal of professionals referred all suspected cases to these specific clinics, using telephone screening or in specific areas of the clinic.<br>• Larger PHC clinics with sufficient supplies (PPE, alcohol, diagnostic test) absorbed the demand of users with influenza but presented a greater risk of leaving affected professionals and difficulties in referring users to the tertiary level. |
| Masotti et al., 2013<sup>16</sup>. (H1N1). | • Set of community-based PHC services, including a gateway to clinical services, home care, and health prevention and promotion programs (eg, smoking).<br>• Restricted role of the federal government in the provision of PHC and public health services, with emphasis on Canadian provinces and territories.<br>• Existence of PHC and public health services with defined functions and different degrees of integration. | • Organization of coping actions must be adapted to different realities: severity of the epidemic; size, location, service capacity and inventory of service supplies; provision of human resources; characteristics of the territory (urban, rural, mixed, presence of traditional populations and vulnerable minorities); and the degree of integration of PHC with public health services; etc.<br>• Operational action plans are essential, detailing the “how to do it” in each situation, and these must be developed with the participation of the largest possible number of actors involved in facing the epidemic.<br>• Planning for situations of reduction in the number of available professionals and unexpected events must be done.<br>• The greater the integration of different actors and services, the greater the effectiveness of the response.<br>• Centralized care services for users with suspected influenza were important to reduce hospital and PHC burden, but the article does not provide detailed results of this experience.<br>• Communication is central, and guidelines and protocols must bring objective and operational messages, preferably from a single source, and appropriate to the different realities.<br>• The safety of professionals and their families was a concern that hampered the integration of several public health actions in PHC and should always be an aspect of attention. |
| Miller et al., 2018<sup>17</sup>.<br>(Ebola). | • Communities with a high rate of viral transmission, up to 2.5 hours away from the nearest PHC service.<br>• Health systems organized differently across the three countries.<br>• Selective community health practices (mostly health care for prevalent maternal and child conditions).<br>• Volunteer CHWs living in the community and working under the supervision of external healthcare workers.<br>• CHWs refer users to PHC services which are of reference for the community.<br>• Poor work structure (insufficient funding, equipment, and medicines with irregular and insufficient provision). | • Stakeholders commented on the provision of community health services before, during, and after the epidemic.<br>• The most effective CHWs in caring for their communities were those receiving timely training, effective and objective information, ongoing support from services and official organizations outside the community, and equipment (including medications).<br>• In places with better structures in the period before the epidemic, responses were more effective during and after the emergency.<br>• In places with greater involvement of local leaders, social mobilization, and time spent by CHWs before the epidemic, the population used and trusted more the services, empowering CHWs as the first contact for people, and improving the reference to PHC services. |
| Phillips et al., 2007<sup>18</sup>.<br>(SARS) | • Physician-centered solo, group, and corporate PHC practices.<br>• These clinics are weakly articulated with governmental governance bodies.<br>• Fragile integration of PHC with public health structures.<br>• Study carried out at a later stage – 6 – of the epidemic. | • Three workflow models were identified for PHC clinics in the epidemic: A. Standard, in which health care continues as usual, including suspected cases; B. “Streamed Model”, in which the clinic makes its own plan to serve only users with influenza, to serve only users with various problems, or to focus on specific services (prenatal, childcare, surgical procedures); C. Mixed, in which each doctor at the clinic draws the list of services that he or she will offer to users, without joint planning, whether or not he or she can assist people with suspected influenza.<br>• Model “b” was considered safer and more effective in this epidemic, although it needs more structural support.<br>• Model “a” was considered more accessible and flexible and may be better accepted by users, but it increases the risk of contagion among healthcare workers and may not offer appropriate care to people in isolation at home.<br>• Public health practices, such as surveillance or home monitoring of confirmed and suspected cases, are particularly difficult to integrate with PHC in Australia but can be more easily adopted in model “b” and, if any healthcare workers plans to do so, in “c”.<br>• All models must be planned according to their installed capacity and profile and number of available professionals, it being important to carry out an adequate planning of the rear of healthcare and administrative workers. |
| Sarikaya, Erbaydar, 2007<sup>19</sup>. (H5N1). | • Community-based PHC with problems of access, structure, healthcare workers provision, and integration with public health services.<br>• PHC chiefly provided via federal government.<br>• Turkey had the first cases outside the Asian and African continents.<br>• Study done in phase 3 of the epidemic. | • Search for PHC increased significantly when the Ministry of Health declared officially the epidemic.<br>• The more effective services had adequate human resources and supply of PPE and other equipment, outlined action plans, greater integration with epidemiological surveillance and good communication strategies with the community.<br>• PHC was not responsible for compulsory notification of diseases until the epidemic.<br>• There was a gradual integration of PHC with epidemiological surveillance throughout the epidemic, including better organization of notification and identification and monitoring of cases by health teams, which played an important role in facing the problem.<br>• The study mentions the importance of telephone counseling services, but there is no more detailed information to allow further consideration.<br>• Part of the qualitative results suggest that the PHC bond and community orientation contribute to the process of identifying cases and guiding people. |
| Siekman et al., 2017<sup>20</sup>. (Ebola). | • A study carried out in the context of the same epidemic analyzed by Miller et al. (2018), but focusing on specific cities in Liberia.<br>• Location with a fragile health system, serious problems of access, longitudinality and provision of human and material resources.<br>• They analyze the performance of voluntary CHWs, living in the community, in the provision of selective health services for the most frequent maternal and child conditions. | • Most CHWs were able to maintain care for the population during the epidemic, sometimes with an increase in consultations.<br>• The scope of action of CHWs varied, including home visits, meetings for health education, treatment of acute diseases such as pneumonia, early identification of Ebola cases, and referral of suspected cases to PHC services.<br>• The determining factors for the positive results were: choice of people from the community itself; timely training and supervision with the development of action protocols; availability of equipment and medicines; external functional PHC services; non-monetary incentives for |
|  | CHWs working with precarious external supervision and reference to health services, in addition to difficult access, insufficient equipment and medicines, and conflicting relationship between users and official health services. | volunteers; and stable project financing.<br>The greatest risk is in the potential for contamination of CHWs. |

CHW: community health workers. PHC, primary health care. PPE, personal protective equipment

### Reality and context in which the service is inserted

As shown by Masotti et al. ^16^, reflecting from the context of an H1N1 epidemic in Canada, there are no universal and linear responses to such a crisis. Many factors must be taken into account in preparing an effective response, including the characteristics of the epidemic itself, the territory, the system, and the health services.

Another fundamental point concerns cultural aspects. Both Miller et al. ^17^ and Siekmans et al. ^20^, studying the same Ebola epidemic that began in 2014 in countries on the African continent, noted that the search for facility-based primary care services by rural communities is influenced not only by the distance and social vulnerability but also by the fear of contracting the disease in the service and by a deep distrust in the official structures as a result of previous armed conflicts. In both cases, the formulation of community-based services with volunteer workers responded not only to the country’s precariousness and scarcity of resources but also contributed to increasing people’s bond with PHC and thus establishing access and the possibility of treatment.

However, the formatting of the health system seemed to be the main factor to be considered when formulating responses to an epidemic. The selected studies are from a few countries, but illustrate some ways of organizing PHC in the world. We found studies carried out in systems that have community-based primary care services relying on multiprofessional work and offering a good diversity of services to the population, including home care and health surveillance ^14,16,19^. We identified studies carried out in systems whose PHC is mostly provided privately by small clinics or offices owned by doctors themselves or belonging to local corporate networks, featuring a fragmented system focused on individual care, with fee-for-service paid trough health insurance or out-of-pocket ^15,18^. In the African context ^17,20^, the studies present a scenario of profound health deficiencies, with PHC services difficult to access to a significant part of the population, subject to frequent interruptions of care due to armed conflicts and discontinuity of financial resources.

No study has compared these different models of organization of the health system in the face of an epidemic, being impossible to conclude definitively for the greater or lesser effectiveness of any of these. However, the data suggests community-based PHC services allow health systems to develop effective and comprehensive responses for populations in situations of epidemic, enabling the interplay of individual clinical care with collective actions of epidemiological surveillance, as well as the possibility of further adjustment of work processes and structure for specific healthcare to suspected cases. Small PHC clinics focused on the physician had less potential for such adjustments, despite the possibility of increasing the service capacity for non-commercial hours. But in these cases, the risk of contagion of professionals is greater and issues related to the provision of protective equipment and medications are more problematic.

These contexts also have profound differences in the articulation of PHC with the other levels of care, with public health services and agencies, and with the other social sectors. Where such integration was more problematic [eg. in Turkey ^19^, PHC was not obliged to notify cases of H5N1], there was a greater likelihood of disorganization of workflows, difficulties in referring users to other services, problems with the adoption of protocols and clinical guidelines and provision of essential supplies. Most of the selected studies emphasize the importance of this intra- and intersectoral articulation, especially between PHC and public health agencies, and PHC with urgent and emergency services. This integration reduces the level of stress to which PHC is subjected in similar situations and allows for more effective and direct action with the affected people and those who need the primary care service for their regular monitoring.

### Planning of communication actions and strategies

The effectiveness of the response to the epidemic also depended on the type of action plan and communication strategies constructed. At the beginning of the epidemic, it was common for health professionals and the population to face deficiency or an excess of information about the problem, coming from the most varied institutions and through the most diverse channels. In general, these studies show that such plans must be as operational as possible, focusing on what is especially necessary to be done in the health service that directly impacts the quality of care offered, with the greatest possible safety for users and professionals ^14–16,19^.

Some studies, especially those from fragmented systems in developed countries ^14–16^, show that healthcare workers can receive contradictory or redundant messages from different sources: their own institutions, local and national health authorities, media and scientific literature. Action plans, protocols, and clinical guidelines must be designed in order to facilitate access to the necessary information that will impact the health care outcome.

Another aspect is that such plans should ideally involve the maximum number of stakeholders directly or indirectly involved in coping with the epidemic: managers, healthcare workers, community leaders, and essential professional and non-governmental organizations can present different perspectives on what needs to be addressed in structuring responses to the crisis.

### Provision of essential equipment

The studies shows that the results of the actions of the primary care services depend on the adequate and permanent provision of essential supplies not only to the user but also for the protection of the healthcare worker. Services that managed to maintain an adequate supply of their stocks showed more favorable results in terms of offering access to essential care.

The high potential for contagion is a central concern for healthcare workers when they are at the forefront, providing care for suspected cases. One of the main aspects of maintaining people’s easy access to care is the workforce having sufficient numbers and satisfactory physical and mental health. These people are under deep stress and experience fear of their own illness and potential death, but mainly they fear for the lives of their family members. In some articles, the healthcare workers were removed from the forefront not only because of their won illness but also for the protection of their closest people. The lack of protective equipment increases the risk to which workers are subject and means serious losses in the service’s potential for healthcare delivery.

Problems with the provision of essential supplies and equipment were greatest in services in Turkey ^19^ and some countries on the African continent ^17,20^, although shortages of medicines and personal protective equipment have occurred in more developed countries as well. Planning for maintaining service stocks at safe levels should be done as soon as possible before the number of cases explodes.

### Bond with the community

The bond of healthcare workers with the community appeared in particular in studies carried out in the most vulnerable countries. Sarikaya and Erbaydar ^19^ show how the relationship previously established by family doctors with their community allowed the identification of the first H5N1 cases in the territory, representing the first step in the articulation of epidemiological surveillance and public health actions. The same authors also show that the PHC workers’ detailed knowledge of people’s social reality was poorly utilized by the health system, wasting a potentially opportunity for reducing some conflicts observed with the community.

Miller et al. ^17^ and Siekmans et al. ^20^, analyzing the contexts of Guinea, Liberia, and Sierra Leone, point out that the selection of community health workers resident in the communities was a decisive factor for the good results observed. The history of armed conflicts and social neglect means that a significant part of people living in rural communities have problematic relationships with state or non-government services, increasing the chance that people will choose traditional health practices available in the community, which can result in diagnostic delay and complications. The bond established by the local health agent increases the effectiveness of the educational practices offered by him and increases access to PHC services.

### Work processes, healthcare flows, and specific services

In general, the authors describe a set of reformulations made by the services: separation of rooms, collective spaces, and entrances for the exclusive care of suspicious people; managing cases in an external environment, such as service parking or even community equipment; reallocation of healthcare workers to exclusively attend symptomatic people and/or to carry out administrative and epidemiological surveillance activities; definition of specific protocols and guidelines for healthcare workers in contact with symptomatic people. Such measures have the potential to streamline the service, increase the workers’ safety, rationalize the use of protective equipment, and reduce the team’s stress level. However, it was not possible to draw large conclusions about the effectiveness of these actions, and healthcare reorganizations might need to be tailored to the local, cultural, and social reality.

In the context of a respiratory disease epidemic, building reference services for affected people is of special interest. Two studies ^15,16^ addressed this issue but did not present enough data to about the effectiveness of these reference services. The authors comment that such services were relevant for reducing the burden of care both at the hospital and the PHC level and that being backed by these services is important for small PHC clinics with reduced equipment and supplies, but they do not provide more information on the rational use of protective equipment and medicines, agility in the referral process, or even the level of quality and adoption of clinical guidelines for these services in comparison with the other levels.

## Discussion

The results of the studies reviewed here suggest that a combination of strategies for organizing healthcare flows, greater integration of healthcare workers and services, effective communication, clear definition of roles and actions of the different actors and enhancing the previously established links between PHC and the territory, may generate the most effective responses of the health system to a major epidemic, like in the several examples of what has been experienced in the world in recent decades and the current pandemic by COVID-19.

The seven articles analyzed also point to the need to not only organize the actions of containment, mitigation, suppression and recovery in the face of a pandemic but to learn about the best alternatives for intervention, both individually and collectively ^9,21^. These studies demonstrate that it is necessary to adopt a more systematic approach to the production of scientific knowledge in the face of these crises, in order to understand what needs to be done in order to establish a more effective and efficient organization of the health system.

Evidently, these crises demand rapid responses in contexts of great uncertainty and urgency ^22^. Therefore, it is essential that good scientific evidence is produced timely for great learning ^23^ and, in our view, this movement should constitute one of the central aspects of action plans in the face of pandemics.

Specifically addressing the Brazilian context, the most appropriate PHC model to provide effective, comprehensive health care during the pandemic might be the Family Health Strategy (FHS), because of its community orientation, multiprofessional teamwork, deep territorial connection and intersectoral integration ^24^. PHC models restricted to small doctor-focused services, which in Brazil are more common in supplementary health ^25^, may be more fragile in these situations due to the high risk of contagion of their professionals and less integration with the structure public health.

Some aspects addressed in the articles could be better studied and implemented in the context of the FHS, in order to enhance their results. We cite the strengthening of Community Health Workers (CHW), not only in numerical terms [coverage by CHW has been reduced recently ^26^] but also in qualitative terms, with adequate training for early identification of cases, guidance of the population regarding healthcare flows, and health education; and with the computerization and incorporation in the daily lives of the teams of communication technologies allowing easy integration and communication with the whole healthcare network, diversification of service options for people, and real-time monitoring of the local epidemiological situation. It is also worth reflecting on the extent to which the National Primary Care Policy (PNAB) adopted in recent years leads to a more powerful PHC in a context of profound social inequalities, underfunding of the health system and high epidemiological complexity ^24,26^.

Another aspect present in two reviewed studies ^15,16^ dialogues with a strategy that appears as an intervention option in the COVID-19 pandemic in several Brazilian municipalities ^27^, which it is the creation of specific services for the care of suspected cases. No study among those analyzed had the central objective of evaluating the effectiveness of these services in the context of major epidemics, which allows us only to hypothesize that there are positive effects of their creation in terms of reducing the burden of care both in PHC and in the urgency and emergency network, and that such a level of centralization can result in a more rational administration of inputs, personal protective equipment, and medications, as well as more effective coordination of the reference of suspected cases between the levels of care.

Some limitations of this work must be considered. Rapid reviews are good options for producing and systematizing knowledge in a timely manner for evidence-based decision making ^12,28,29^ at the cost of some flexibility in essential steps of systematic reviews. We chose to qualify some steps recommended for quick reviews, which increases its internal validity: we searched three databases of impact; we kept two independent researchers in the screening stages, selecting the corpus of articles for narrative review, and extracting the data; time limits and geography were not adopted in the selection of studies; and we incorporated articles in more than one language. The main limitations in this study are the lack of individualized analysis of the quality of the selected articles and the failure to carry out manual searches and gray literature, which may have excluded from the review relevant papers available in institutional technical reports or papers not published in indexed journals.

In summary, this review points to the need to build action plans with broad participation by the actors involved in coping with the epidemic, whose determinations consider the context in which the service is inserted, condensing work process adjustments and structuring healthcare flows, diversification of care practices and integration with the whole of the care network, especially with the public health apparatus. Emphasis is placed on the importance of empowering the service link with its registered population.

Unfortunately, such measures do not fit into proposals to reduce the participation of public authorities in guaranteeing the constitutional right to health.

## Data Availability

This is a rapid systematic review. All relevant data are described in the article.

